# Pre- and Post-synapses Contain Lecanemab-reactive Amyloid-β in Post-mortem Human Alzheimer’s Disease Brain

**DOI:** 10.64898/2026.05.08.26352549

**Authors:** Kristjan Holt, Ya Yin Chang, Mosi Li, Giulia Albertini, Colin Smith, Jane Tulloch, Bart De Strooper, Giles E. Hardingham, Tara L. Spires-Jones

## Abstract

Recently, the amyloid-beta (Aβ) targeting antibody lecanemab has demonstrated modest therapeutic efficacy in slowing cognitive decline in people with Alzheimer’s disease (AD). Lecanemab clears amyloid plaques from the brain; however, plaque load does not correlate strongly with cognitive function. The strongest neuropathological correlate of cognitive decline in AD is synapse loss, which is exacerbated in the halo surrounding neuritic amyloid plaques where Aβ accumulates in remaining synapses. Here, we hypothesised that, through clearing plaques and the associated halo of soluble Aβ that can directly damage synapses, lecanemab could temper plaque-associated synapse loss. High-resolution imaging of temporal cortex tissue from people who died with AD (N=20) and age-matched controls (N=19) reveals lecanemab staining within individual pre and post-synaptic excitatory terminals in addition to plaque staining. The percentage of pre-synapses containing lecanemab-positive Aβ was over 200% higher in AD and the percentage of post-synapses was over 150% higher in AD than control tissue, with highest levels of synaptic lecanemab staining observed near plaques. These data demonstrate that lecanemab antibody recognises Aβ within synapses, warranting future work to determine whether lecanemab treatment slows cognitive decline, at least in part, through both clearing plaques and facilitating clearance or neutralisation of synaptic Aβ.

## Introduction

Lecanemab, a humanized IgG1 monoclonal antibody derived from preclinical murine mAb158, has demonstrated clinical efficacy in modestly slowing cognitive decline in people with Alzheimer’s Disease (AD)^1,2^. Lecanemab binds soluble Aβ with high affinity—in particular, protofibrils—and effectively clears A® in the form of extracellular plaques from the brain^1^. As such, the ability for lecanemab to abate cognitive decline is likely linked mechanistically to its capacity to bind and remove Aβ from the brain. However, critical to note is that a clearance of plaques alone from the brain fails to explain the clinical efficacy of lecanemab. When compared against other neuropathological features of AD, plaque burden represents a comparatively poor correlate of clinical symptoms^3^. Furthermore, significant plaque deposition is frequently reported upon post-mortem examination of individuals who died with no evident dementia during life^4^. Hence, it is likely that lecanemab’s efficacy lies not solely in its capacity to clear plaques *per se* but perhaps also through an ability to indirectly modulate other pathological processes by removing upstream toxic Aβ.

Region-specific loss of synapses is the strongest pathological correlate of declining cognition in AD^3,5,6^. Profoundly reduced synapse density is often observed within the vicinity of densecored neuritic plaques; correlating spatially with a high concentration of soluble Aβ oligomers that are understood from model systems to be highly synaptotoxic^7,8^. In mouse models, passive immunotherapy with an antibody recognizing Aβ allows synapse recovery around plaques even when plauqes remain present ^9^, indicating binding of soluble Aβ is sufficient to redeuce synaptotoxicity. We previously observed intrasynaptic Aβ around amyloid plaques in human AD brain tissue with multiple antibodies including those recognizing both small and large oligomers ^8,10–12^, together leading to the supposition that plaque-associated synaptic loss may occur as a consequence of direct toxicity incurred by molecular interaction of synaptic components with Aβ oligomers. However, it was previously unknown whether the therapeutic antibody lecanemab binds synaptic Aβ. We hypothesized that lecanemab antibody would recognize Aß both within plaques and within individual synapses around plaques, which could have implications for the therapeutic activity of the antibody. To test this, we used post-mortem brain tissue samples examined with single-synapse resolution array tomography (AT) and immuno-electron microscopy.

## Materials and Methods

For full details of all experimental procedures/materials, refer to **Supplementary Methods**.

### Human Cases

#### Ethical Approval and Case Demographics

Post mortem-samples were acquired from inferior temporal gyrus (Broadmann Area 20/21). Case selection was based on clinical and neuropathological diagnoses. Inclusion and exclusion criteria are detailed fully in **Supplementary Methods**. The final number of cases for this study included *N = 20* AD (9 males; 11 females) and *N = 19* Control (10 males; 9 females), with groups matched for sex, age (AD: 82.3 ± 7.8 years, Control: 81.3 ± 2.8 years) and postmortem interval (PMI). Study demographics are included in **Table 1** with each case denoted by an anonymised identifier (BBN number). Ethical approval for the use of human post-mortem tissue was reviewed and approved by the Edinburgh Brain Bank Ethics committee (No.: 11/ES/0022) and the ACCORD medical research ethics committee (AMREC), a joint office of the University of Edinburgh and NHS Lothian (No.: 15-HV-016).

**Table 1:**
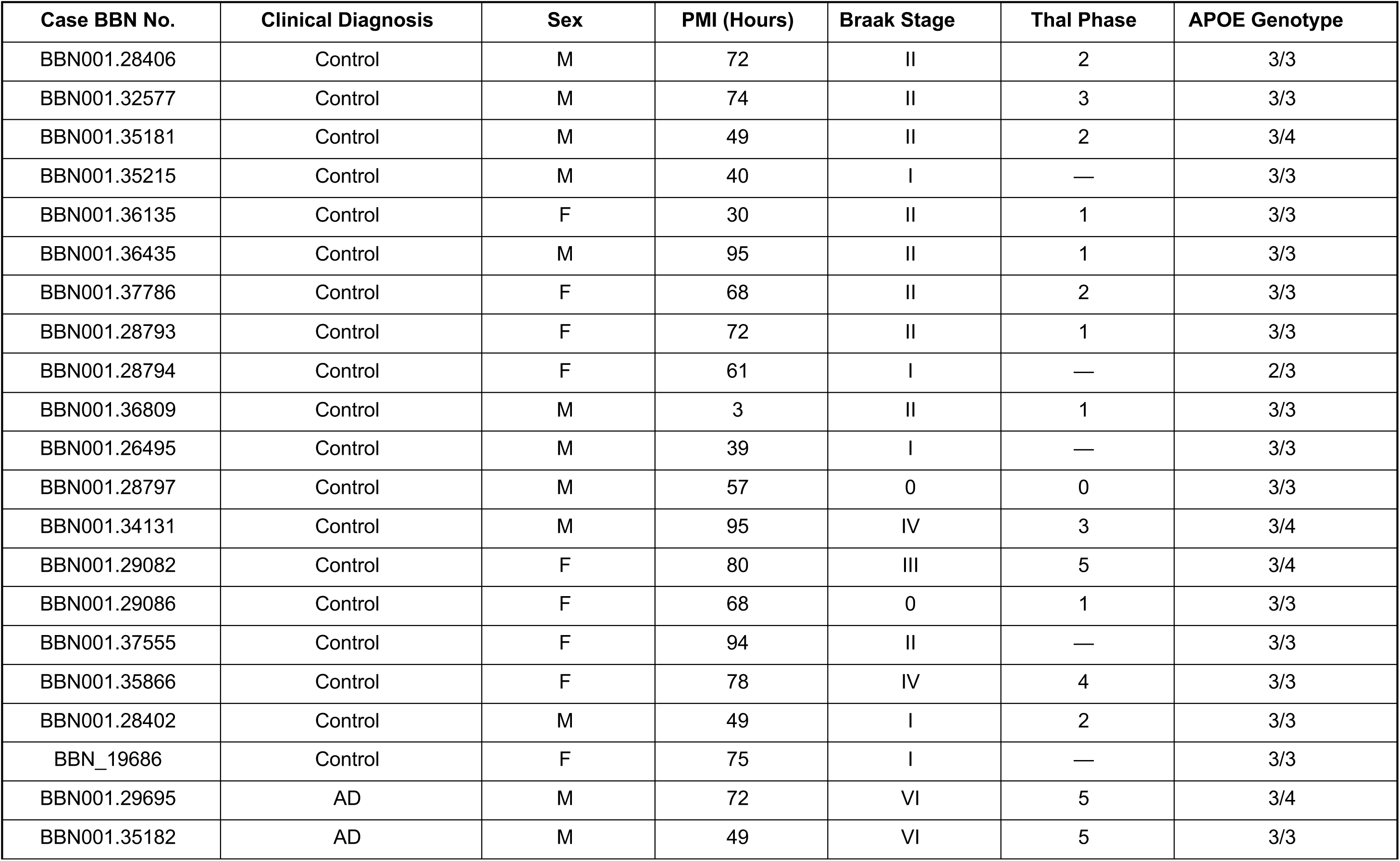
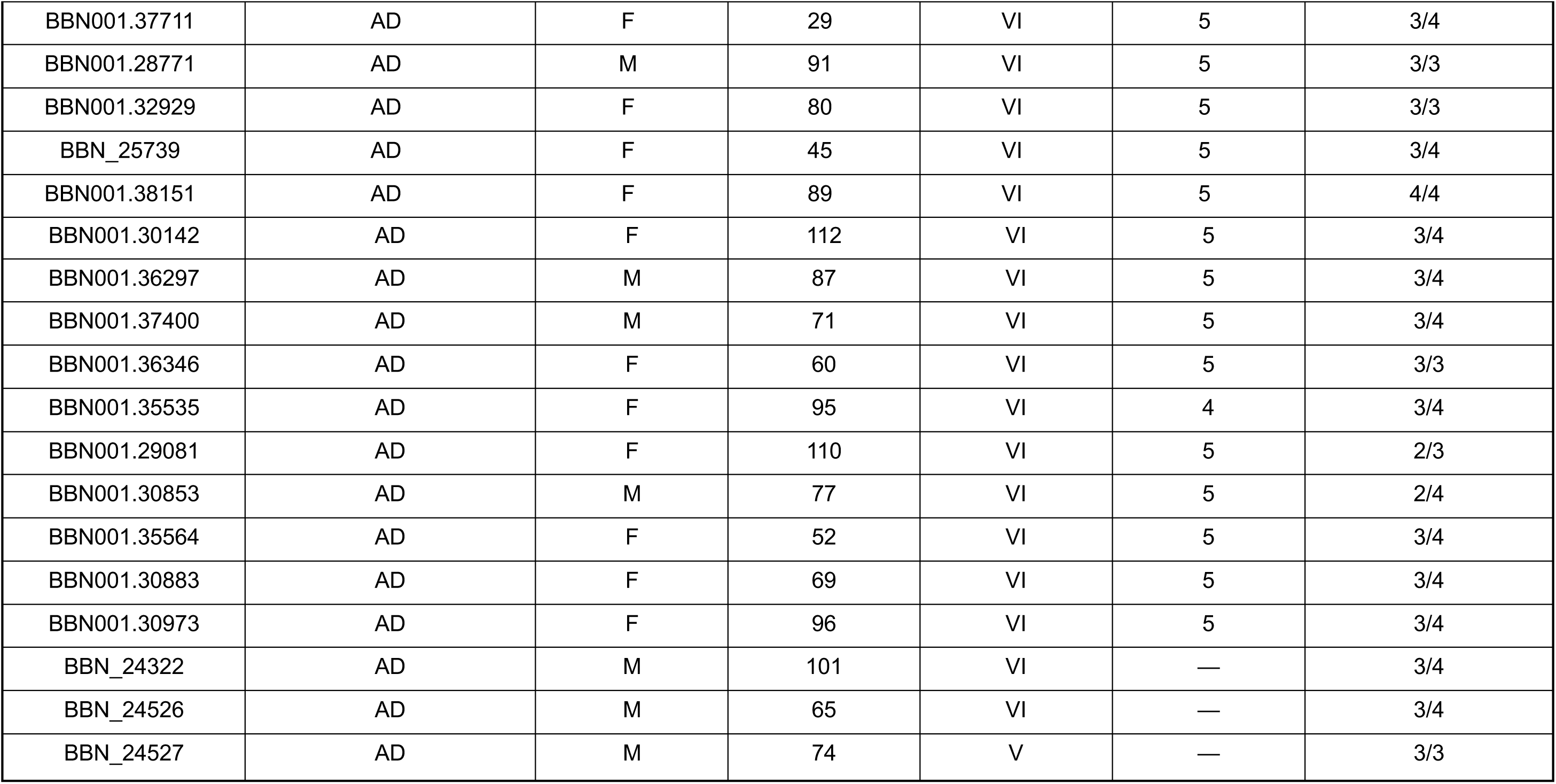
Full demographic details of cases included in this study, including clinical diagnosis, sex, age, postmortem interval (PMI), Braak Stage, Thal Phase and APOE genotype.

| Case BBN No. | Clinical Diagnosis | Sex | PMI (Hours) | Braak Stage | Thal Phase | APOE Genotype |
| --- | --- | --- | --- | --- | --- | --- |
| BBN001.28406 | Control | M | 72 | II | 2 | 3/3 |
| BBN001.32577 | Control | M | 74 | II | 3 | 3/3 |
| BBN001.35181 | Control | M | 49 | II | 2 | 3/4 |
| BBN001.35215 | Control | M | 40 | I | — | 3/3 |
| BBN001.36135 | Control | F | 30 | II | 1 | 3/3 |
| BBN001.36435 | Control | M | 95 | II | 1 | 3/3 |
| BBN001.37786 | Control | F | 68 | II | 2 | 3/3 |
| BBN001.28793 | Control | F | 72 | II | 1 | 3/3 |
| BBN001.28794 | Control | F | 61 | I | — | 2/3 |
| BBN001.36809 | Control | M | 3 | II | 1 | 3/3 |
| BBN001.26495 | Control | M | 39 | I | — | 3/3 |
| BBN001.28797 | Control | M | 57 | 0 | 0 | 3/3 |
| BBN001.34131 | Control | M | 95 | IV | 3 | 3/4 |
| BBN001.29082 | Control | F | 80 | III | 5 | 3/4 |
| BBN001.29086 | Control | F | 68 | 0 | 1 | 3/3 |
| BBN001.37555 | Control | F | 94 | II | — | 3/3 |
| BBN001.35866 | Control | F | 78 | IV | 4 | 3/3 |
| BBN001.28402 | Control | M | 49 | I | 2 | 3/3 |
| BBN_19686 | Control | F | 75 | I | — | 3/3 |
| BBN001.29695 | AD | M | 72 | VI | 5 | 3/4 |
| BBN001.35182 | AD | M | 49 | VI | 5 | 3/3 |
| BBN001.37711 | AD | F | 29 | VI | 5 | 3/4 |
| BBN001.28771 | AD | M | 91 | VI | 5 | 3/3 |
| BBN001.32929 | AD | F | 80 | VI | 5 | 3/3 |
| BBN_25739 | AD | F | 45 | VI | 5 | 3/4 |
| BBN001.38151 | AD | F | 89 | VI | 5 | 4/4 |
| BBN001.30142 | AD | F | 112 | VI | 5 | 3/4 |
| BBN001.36297 | AD | M | 87 | VI | 5 | 3/4 |
| BBN001.37400 | AD | M | 71 | VI | 5 | 3/4 |
| BBN001.36346 | AD | F | 60 | VI | 5 | 3/3 |
| BBN001.35535 | AD | F | 95 | VI | 4 | 3/4 |
| BBN001.29081 | AD | F | 110 | VI | 5 | 2/3 |
| BBN001.30853 | AD | M | 77 | VI | 5 | 2/4 |
| BBN001.35564 | AD | F | 52 | VI | 5 | 3/4 |
| BBN001.30883 | AD | F | 69 | VI | 5 | 3/4 |
| BBN001.30973 | AD | F | 96 | VI | 5 | 3/4 |
| BBN_24322 | AD | M | 101 | VI | — | 3/4 |
| BBN_24526 | AD | M | 65 | VI | — | 3/4 |
| BBN_24527 | AD | M | 74 | V | — | 3/3 |

### Processing, Immunostaining and Imaging of Human Tissue

#### Array Tomography

Lecanemab antibody was provided by co-authors Giulia Albertini and Bart De Strooper. This antibody has been characterized previously in Albertini *et al.*, Nature Neuroscience, 2025^13^. Array tomography was performed in line with protocols described previously^14^. Briefly, tissue was collected at autopsy, fixed in paraformaldehyde, dehydrated, and embedded in LR white acrylic resin. Samples were cut into ribbons of 70 nm ultrathin sections and immunostained using primary and secondary antibodies, details of which are included in **Supplementary Table S1**. Images of serial sections were acquired at 63x magnification and processed for downstream analysis using an in-house pipeline. Full details of immunostaining and image analysis, including links to custom analysis scripts, are detailed in **Supplementary Methods**.

#### Immunogold Electron Microscopy (iEM)

Immunogold processing, staining and imaging were performed as described previously^14^ ^15^, full details of which can be found in **Supplementary Methods**. Briefly, Resin-embedded tissue blocks were sliced into 50.0nm ultrathin sections and mounted onto formvar/carbon-coated nickel grids, Sections were incubated with lecanemab primary antibody and a 10 nm gold-conjugated secondary antibody (**Supplementary Table S1**).

### Statistical Analysis

All statistical analyses were performed using *R* (version 4.2.1, June 2022) via linear mixed effects modelling or standard linear models as detailed in supplementary methods. Data were assessed for normality via visual inspection of histograms and Shapiro-Wilk tests. Models were adjusted for sex, postmortem interval and APOE4 status with tissue block nested within case fitted as a random effect to account for within-subject variance. Model fit was assessed by examination of residual plots conjunct with formal fit testing and data transformed where required. Post-hoc tests were adjusted for multiple comparisons via Holm’s correction. For analysis of synapses as a function of plaque distance, data found to be normally distributed were assessed via linear models accounting for sex, postmortem interval and APOE4 status, or alternatively, unpaired two-sample Wilcoxon rank sum test.

In both cases, data from each binned distance (10-50µm) were compared against the average (mean or median, respectively) of plaque-absent image stacks from the control group and *p-values* adjusted via Holm’s posthoc test for all five comparisons.

## Results

### Lecanemab Antibody Labels both Dense Core and Diffuse Plaques

Previous data has demonstrated that in postmortem human AD temporal cortex, lecanemab antibody positively labels amyloid plaques with conventional immunohistochemical staining^16^. We confirm using high-resolution array tomography that lecanemab antibody labels both dense core and diffuse plaques, and observe that plaques are surrounded by a halo of small lecanemab positive deposits as we have observed previously with other Aβ antibodies **(Figure 1A-B)**^8^ ^12^ ^11^. Image stacks were acquired in areas containing plaques and regions distant from plaques in each case. Although plaques were rarer in our control samples, over half of controls (11/19 — 57.9%) had at least one plaque **(Table 1)**. As expected, quantification of these image stacks demonstrated the burden of lecanemab (calculated as percent volume occupied by immunostaining) was higher in regions with plaques than in regions without **(Figure 1C)**. Linear mixed effects modelling of the data revealed a significant main effect of plaque presence (F(1, 184.599) = 218.89, *p < 2e-16\*\*\**). In control cases, quantifiable lecanemab signal was still evident, particularly where plaques were present. Despite balancing control and AD sampling of plaques to focus on plaque-associated changes, there was a significant main effect of diagnosis (F(1, 33.198) = 4.21, *p* = 0.048*); reflecting an increase in median plaque volume in AD cases over controls, and an increase in lecanemab signal volume when comparing imaging fields in which no plaque was present between groups. However, due to the highresolution nature of array tomography which is designed to study small structures like synapses, very few plaques were sampled from each case (*N* = 2-3 plaques on average; **Supplementary Table S2)**. To confirm that lecanemab immunofluorescence signal observed with array tomography reflected a genuine binding of this antibody to plaques, we used immuno-electron microscopy. Ultrastructural imaging revealed lecanemab detected with a gold-conjugated secondary antibody indeed decorated plaque fibrils (**Figure 1D**). Altogether, these data validated that the lecanemab antibody used here binds Aβ, both within plaques and in the surrounding neuropil.

**Figure 1:**
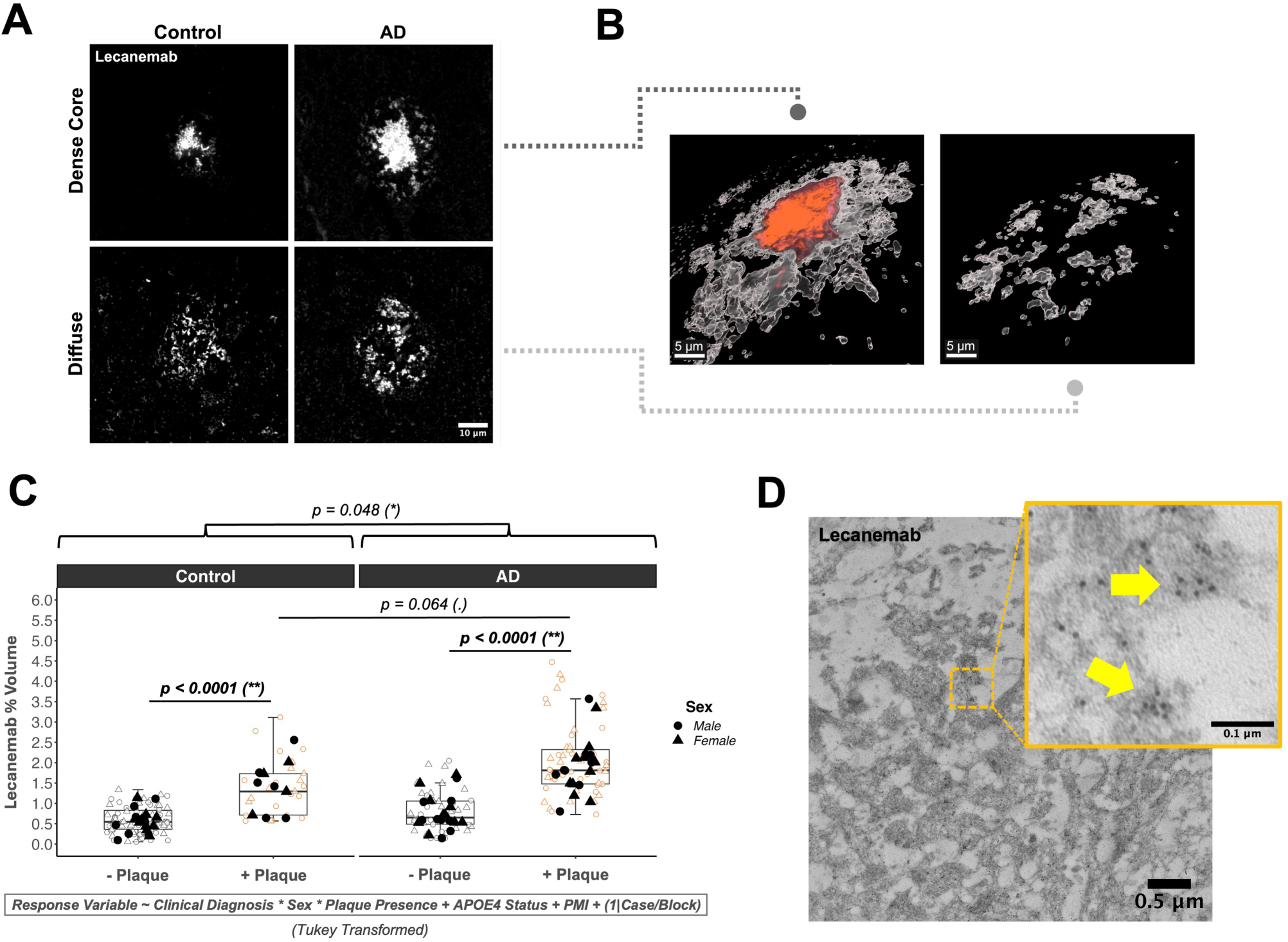
Lecanemab Antibody Labels Plaques and Associated Aβ Deposits. (**A**) Lecanemab positively labels both dense core and diffuse plaques upon AT immunostaining, as profiled across control (*N =* 19) and AD (*N =* 20) cases. (**B**) Three-dimensional reconstructions of a dense core and diffuse plaque from a single AD case reveals the presence of an evident core (orange) and associated halo (white). Diffuse plaques meanwhile lack these features, while exhibiting an amorphous boundary with the surrounding parenchyma. (**C**) Quantification of % lecanemab volume stratified by plaque presence (dense core + diffuse) indicates a trend toward increased plaque size in AD cases. Additionally, the presence of a plaque in the imaging field was associated with significantly increased lecanemab signal volume, as expected. Filled datapoints indicate average (mean) paired synapse density per human case, while clear datapoints indicate underlying technical replicates in the form of individual image stacks (N = 3-6 stacks per case). (**D**) Cross-validation of AT staining via iEM reveals a positive binding of lecanemab to fibrillar A® associated with a nearby plaque (yellow arrows). **Image Sources:** A — single 70nm slices from a serially imaged AT ribbon (63X magnification). As diffuse plaques were dimmer than dense core counterparts, different contrast values were used to highlight them for representative purposes. The same contrast values were used for comparing plaques across Control and AD groups. B — three-dimensional reconstruction of plaques generated using IMARIS software version 10.2.0. **Abbreviations**: Aβ = amyloid-beta; AT = array tomography AD = Alzheimer’s Disease; nm = nanometer.

### Lecanemab Recognises Synaptic Aβ

We next examined whether lecanemab signal is observed in excitatory synapses in the AD brain. Individual synapses were defined as a pre-synaptic object (synaptophysin+) and a postsynaptic object (PSD-95+) located within a maximum distance of 0.5µm of one-another **(Figure 2A)**. Non-paired synaptic staining was excluded from analysis to reduce non-specific noise. The percentage (%) of total paired pre- and postsynaptic puncta colocalising with lecanemab was quantified across all image stacks, including those acquired both with a plaque centred and those distal to plaques. ANOVA analysis of linear mixed effects models including sex as a covariate and adjusting for APOE4 status and PMI revealed a significant main effect of AD diagnosis for both presynapses (F(1, 32.55) = 8.14, *p* = 0.0075**) and postsynapses colocalised with lecanemab (F(1, 32.74) = 10.04, *p* = 0.0033**), reflecting an increase in lecanemab-positive synaptic A® in the AD brain **(Figure 2B-C)**. We observed a 208.82% relative median increase (+0.38% absolute) in presynapses with lecanemab in AD over controls and a 189.59% relative increase (+0.33% absolute) in postsynapses. This was mirrored by an AD-associated 27.79% reduction in mean paired synapse density within the same brain region, as captured by a significant main effect of diagnosis (F(1, 32.61) = 31.24, *p* = 3.37e-06***) **(Figure 2D)**. Additional analysis that sub-stratified experimental groups on the basis of plaque presence in the imaging field indicated that, where plaques were present in the AD group, both pre- and postsynapses were still further likely to be found in association with lecanemab, coupled with a plaque-associated loss of paired synapse density **(Supplementary Figures S1-S2)**. Overall, these data confirmed that, in a brain region with synapse loss, lecanemab antibody recognised Aβ at synapses; did so with greater frequency in the AD brain, and was more likely to be found at synapses near plaques.

**Figure 2:**
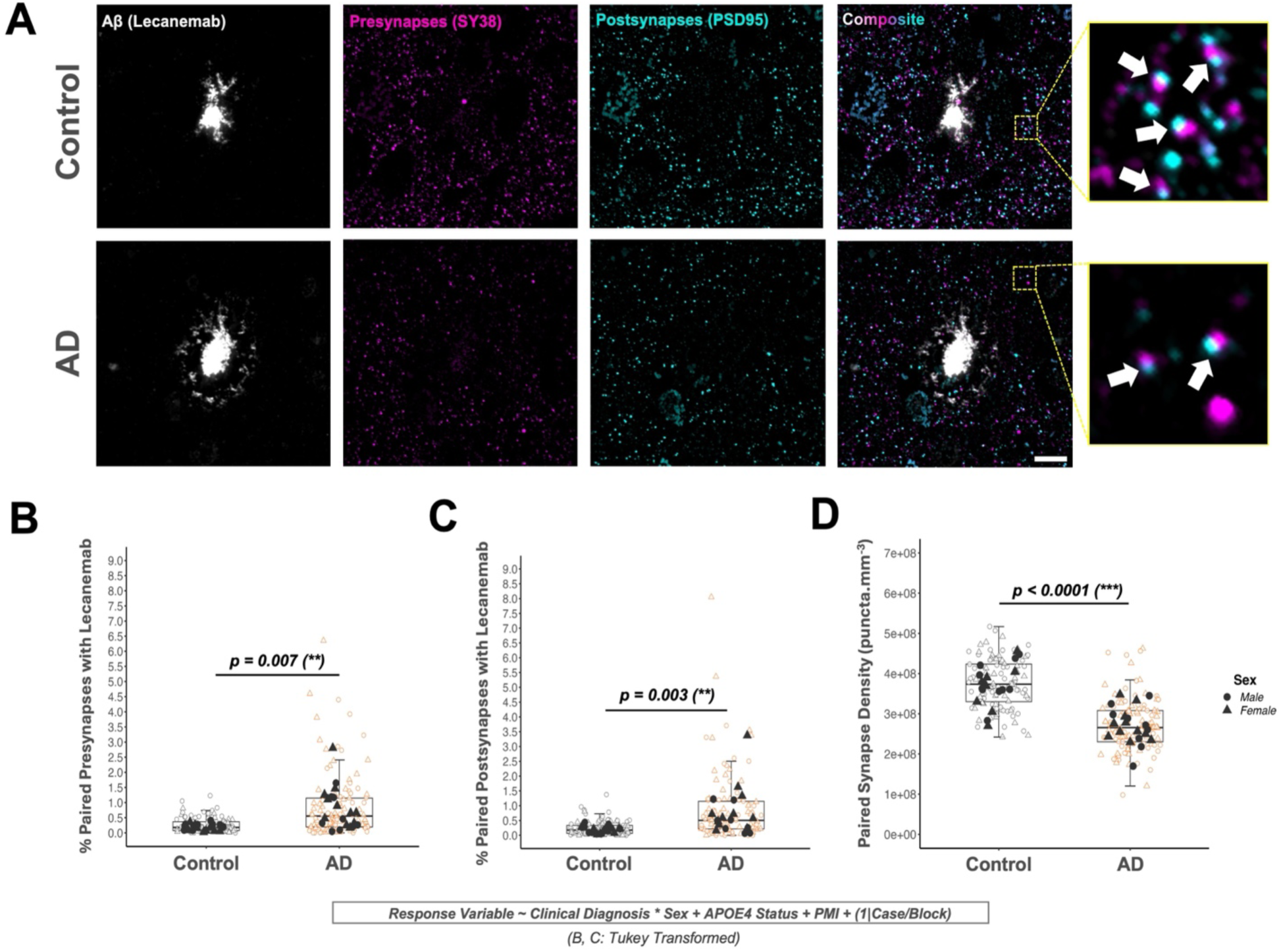
Lecanemab Signal Colocalises with Pre- and Postsynapses and Increases in the AD Cortex. (**A**) Representative images are shown illustrating lecanemab-positive plaques from both control and AD groups, co-labelled with markers corresponding to presynaptic (SY38/synaptophysin; magenta) and excitatory postsynaptic (PSD-95; cyan) puncta. Insets demonstrate “paired” synapses, defined by a presynaptic terminal in direct apposition to a corresponding postsynaptic partner within a distance of 0.5µm (arrows). Only paired synapses were included for analysis. (**B-C**) Quantification of the percentage (%) of paired pre- and postsynaptic puncta found to colocalise with lecanemab reveals that, in the AD cortex, synapses more frequently associate with lecanemab antibody signal. Filled datapoints indicate average (median) synapses with lecanemab per human case, while clear datapoints indicate underlying technical replicates in the form of individual image stacks (N = 3-6 stacks per case). (**D**) Quantification of paired synapse density within the same Z-stacks demonstrates that increased lecanemab-synapse interaction is paralleled by an inverse pattern of synapse loss in AD. Filled datapoints indicate average (mean) paired synapse density per human case, while clear datapoints indicate underlying technical replicates in the form of individual image stacks (N = 3-6 stacks per case). **Image Sources:** A — single 70nm slices from a serially imaged AT ribbon (63X magnification). Scale bar = 10µm. Abbreviations: AD = Alzheimer’s Disease; AT = array tomography; PSD-95 = Postsynaptic Density Protein 95; nm = nanometer.

### Lecanemab Colocalises with Synapses Near Plaques

Having established that quantifiable lecanemab antibody signal could be observed at synapses and further increased where plaques were present, we assessed whether said signal exhibited any spatial correlation with plaque-associated synapse loss more specifically within the plaque halo. Data were filtered to include only plaques with a dense core morphology (*N = 40*) from AD cases (*N = 18*) and, following previously described methods^8,17^, the colocalisation of synaptic puncta with lecanemab was quantified within 10-micron bins extending from the boundary of the plaque core. Lecanemab staining was present in both paired pre- and postsynaptic puncta in the plaque halo as expected **(Figure 3A)**, with those surviving within a radius of 10-20 microns exhibiting significantly increased likelihood of colocalisation. Synapses within more distal bins exhibited less association with lecanemab; rapidly decreasing toward the control baseline **(Figure 3B-C)**. Paired synapse density demonstrated an inverse decrease within the same initial 10-20 micron area near plaques when compared to controls, reflecting plaque-associated synapse loss as reported previously in the literature **(Figure 3D)**. As such, when examining the dense core plaque halo in the AD cortex, synapses that had survived profound loss in the immediate proximity of the fibrillar core, and likely exposed to higher concentrations of synaptotoxic Aβ, appeared more likely to associate with lecanemab when compared against those residing more distally.

**Figure 3:**
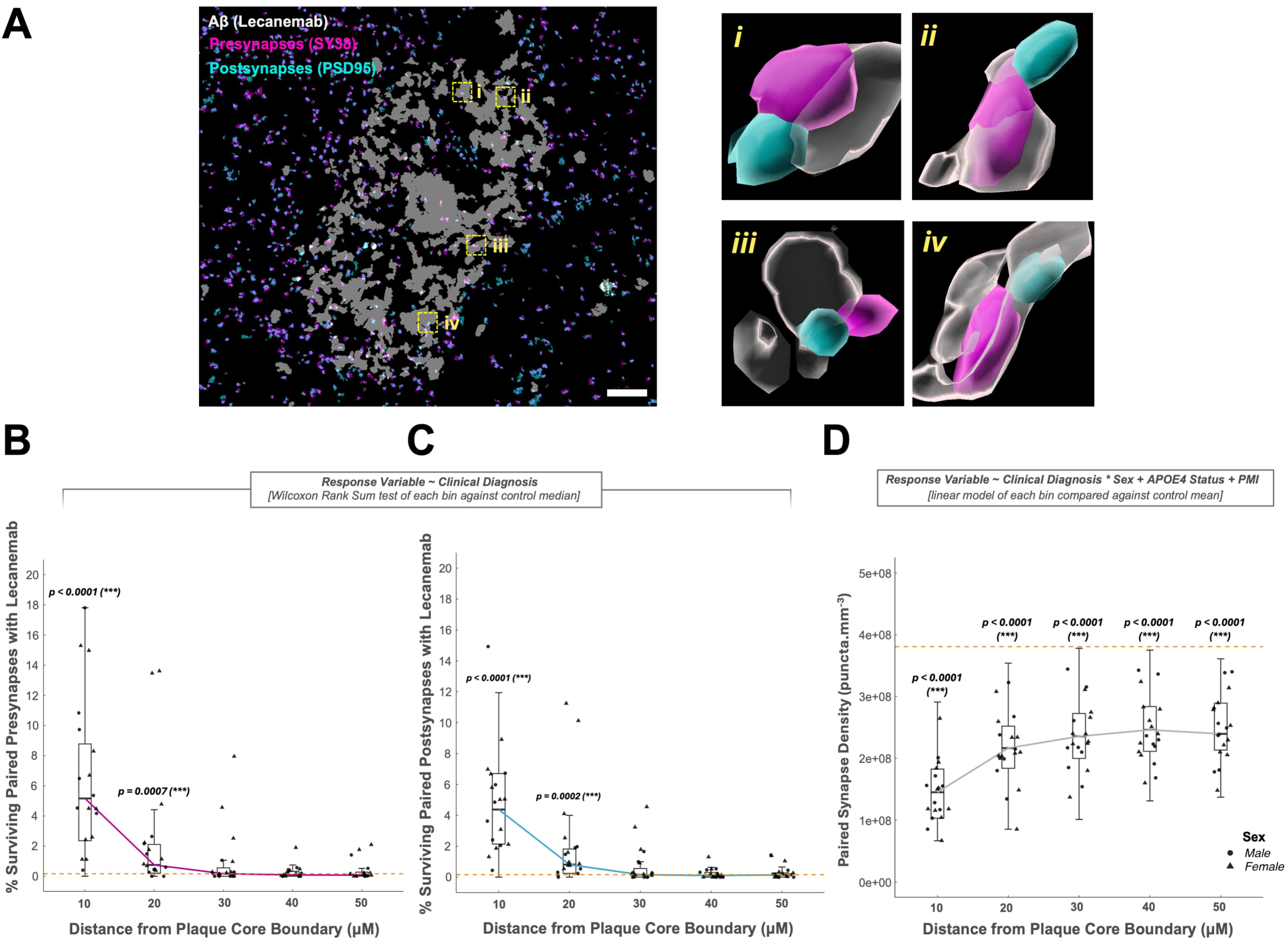
Lecanemab Preferentially Colocalises with Synapses within the Synaptotoxic Dense Core Plaque Halo. (**A**) A maximum intensity projection of a segmented Z-stack corresponding to a dense core plaque from a single AD case is shown with surrounding presynaptic (SY38/synaptophysin; magenta) and excitatory postsynaptic (PSD-95; cyan) puncta. Insets (i-iv) demonstrate a selection of synapses registered as colocalised with lecanemab antibody; all of which are present within the halo surrounding the central core. (**B-C**) Filtering data to include only dense core plaques from AD cases and quantifying the percentage of synapses in binned regions around the plaque in 10µm increments associated with lecanemab from the core boundary, surviving synapses within the immediate 10-20 micron radius proximal to the core exhibit significantly increased likelihood of colocalising with lecanemab when compared against synapses residing within more distal bins. Datapoints represent average values (medians) for each independent case, while the dashed line represents the average (median) value from control Z-stacks with no plaque present. (**D**) Quantification of paired synapse density in the same Z-stacks as a function of plaque distance indicates that the region of increased lecanemab-synapse colocalisation spatially overlaps with profound synapse loss within a 10-20µm proximity to the central core, suggesting that lecanemab is more likely to associate with “at risk” synapses that persist as survivors within the toxic plaque halo. Datapoints represent average values (means) for each independent case, while the dashed line represents the average (mean) value from control z-stacks with no plaque present. **Image Sources:** A — Left: maximum intensity projection generated from a single Z-stack containing a dense core plaque. Images are shown in segmented/binary thresholded format. Scale bar = 10µm. Right: three-dimensional reconstructions of paired pre- and postsynaptic puncta observed to colocalise with lecanemab within the plaque halo. **Abbreviations:** AD = Alzheimer’s Disease; PSD-95 = Postsynaptic Density Protein 95; nm = nanometer.

## Discussion

Here, we report that lecanemab antibody recognises Aβ in both pre-and postsynaptic puncta in human AD brain using high-resolution imaging. Further, the relative proportion of synapses colocalised with lecanemab was increased in AD cases and was further increased in synapses near plaques. How these observations may in turn relate to the therapeutic benefits associated with lecanemab in AD remain an area of active investigation. Soluble oligomeric forms of Aβ which are prevalent around amyloid plaques have been shown to impact synaptic structure and function in model systems including reducing long-term potentiation, enhancing long-term depression, increasing calcium influx, and causing synapse loss ^18^. Direct interactions of Aβ with synaptic proteins including excitatory neurotransmitter receptors, cellular prion protein, and the sigma-2 receptor have all been shown to cause synaptic dysfunction and loss in mouse brain and neuronal culture systems^18^. As such, it could be speculated that therapeutic antibodies, such as lecanemab, through binding to Aβ at synapses, might act to preserve synaptic function by effectively sequestering or otherwise limiting the ability of Aβ to interact with synaptic machinery and provoke synaptic plasticity deficits or glutamatergic excitotoxicity. In support of this idea, passive immunotherapy with multiple Aβ antibodies prevents synaptic plasticity deficits and synapse loss in several mouse models ^9,19–22^. In addition, strong evidence implicates microglia both in Aβ mediated synapse removal and in driving clearance of plaques by anti-amyloid immunotherapy treatment. In mouse models, Aβ “tags” synapses around plaques for microglial phagocytosis ^23–25^. Further, we have observed synaptic material inside microglia around plaques in human AD brain and observed that human primary microglia phagocytose synapses from AD brain more than those from control brain ^26^. In a mouse model with plaques that received xenotransplanted human microglia, lecanemab treatment reduces both plaque pathology and associated neurite damage only when microglia are present and when the lecanemab is able to bind microglia through the Fc fragment, highlighting the importance of microglia in both plaque clearance and downstream reductions in toxicity ^13^. A putative preservation of synaptic function either through direct neutralisation of toxic Aβ or through microglia-mediated clearance, could conceivably contribute to slowing of cognitive decline when scaled up to the whole brain level. It will be of interest in future to examine brain tissue from people treated with lecanemab to determine whether treatment reduces plaque-associated synapse loss or affects microglial engulfment of synapses.

Limitations of this study include the direct application of antibody to fixed, postmortem tissue that has been subject to fixation and permeabilisation. As such, while lecanemab recognizes synaptic Aβ within this experimental context, it is not clear whether the antibody can enter synapses in living people receiving lecanemab treatment. Second, interpretation is limited by the fact that the array tomography imaging method cannot distinguish between intra- and perisynaptic Aβ within the plaque halo. Finally, the experimental methods employed here are ultimately descriptive in nature and cannot provide direct mechanistic insight into the therapeutic actions of lecanemab. Despite these limitations, our finding that lecanemab positive Aβ associates with synapses in human AD leads to testable hypotheses about whether lecanemab treatment might exert some beneficial effects through clearing dysfunctional synapses or lowering synaptotoxic Aβ levels as a consequence of plaque removal.

## Data availability

All spreadsheets and analysis scripts from the final accepted manuscript will be available on Edinburgh DataShare (https://datashare.ed.ac.uk/handle/10283/3076 DOI for these data to be added after peer review). Raw images are available from the corresponding author.

## Acknowledgements

We acknowledge the Edinburgh Neuroscience and FENS-Kavli Network of Excellence communities for fostering a collaborative environment and discussions of the work. We would like to thank the brain tissue donors and their families who make this work possible. Thanks also to Alzheimer Scotland both for funding the brain and tissue bank and for facilitating inspiring input from people with lived experience of Alzheimer’sdisease.

## Supplementary material

Supplementary material, including full methods and additional data to accompany this report is available at *Brain* online.

## Supplementary Materials and Methods

### Ethical Approval and Human Cases

#### Ethical Approval for the Experimental Use of Human Brain Tissue

Retrieval of post-mortem tissue complied with the Human Tissue (Scotland) Act 2006. All experiments involving post-mortem tissue of human origin were performed in accordance with those reviewed and approved by the Edinburgh Brain Bank ethics committee and the ACCORD medical research ethics committee (AMREC). ACCORD is the Academic and Clinical Central office for Research and Development, a joint office of the University of Edinburgh and NHS Lothian (Approval No.: 15-HV-016). The Edinburgh Brain Bank is a Medical Research Council (MRC) funded facility with research ethics committee approval (No.: 11/ES/0022). At no point during this work were confidential or personable details of human brain tissue donors accessible to experimenters. All data were acquired and processed in a pseudonymized format that excluded all personally identifying characteristics from post-mortem samples. Experimenters had access only to characteristics that were relevant to the research being conducted, including: donor age, sex, APOE genotype, postmortem interval (PMI), neuropathological evaluation (Braak stage/Thal phase), brain weight and brain pH.

#### Human Case Selection and Study Demographics

Selection of human cases was performed on the basis of both clinical and neuropathological diagnoses. All tissue was sampled from the neocortex; derived from the middle-inferior temporal gyrus (Brodmann Area 20/21; “BA20/21”). For cases to be positively classified as having AD, inclusion criteria dictated both a pre-mortem clinical diagnosis of dementia in addition to post-mortem confirmation of AD hallmarks (both A® and pTau) via neuropathological staging (tau Braak V-VI). Conversely, for a case to be classified as a “control”, inclusion criteria were absence of an AD clinical diagnosis and confirmed low to moderate levels of AD neuropathological hallmarks upon post-mortem evaluation of brain tissue (Braak 0-IV). Exclusion criteria included either evidence of major haemorrhage or confirmed presence of substantial amounts of secondary pathology (e.g. Lewy Body inclusions) within the brain region investigated. Furthermore, cases were excluded in the event that post-mortem neuropathological assessment reported evidence of hallmarks leading to a conclusion of other neurodegenerative diseases (e.g. Vascular Dementia or Progressive Supranuclear Palsy). Data regarding clinical diagnosis and neuropathology used to define inclusion and exclusion criteria were obtained from clinical charts provided by the Edinburgh Brain Bank (EBB). The final number of cases included for study encompassed *N = 20* AD (9 males:11 females) and *N = 19* Control (10 males:9 females). All cases were matched on the basis of sex, age and postmortem interval (PMI); however, the APOE4 genotype was significantly overrepresented in the AD group as opposed to APOE3, which was more common in controls (Chi-Squared Test: *p* = 0.002**). Any confound arising from differences in APOE4 status was adjusted for via inclusion of this variable as a fixed effect covariate in downstream statistical analysis.

### Synthesis of Lecanemab mAβ for Immunostaining

Monoclonal lecanemab (0.7mg/mL stock concentration) was acquired courtesy of Professor Bart De Strooper and Dr Giulia Albertini; synthesis of which was performed in accordance with the protocols detailed in Fertan *et al.*, 2025^1^. Briefly, lecanemab was generated on a human IgG1 backbone using publicly available variable domain sequences. This antibody has been validated *in vitro* to bind synthetically-generated Aβ_1-42_ and pE-Aβ_3-42_ (pyroglutaminated N-truncated Aβ) co-aggregates with high affinity in a concentration-dependent manner, as determined by single-molecule array (SiMoA) scan; details of which can be found in the results published previously by Fertan *et al.*, 2025^1^.

### Processing of Human Brain Tissue

#### Tissue Processing for Array Tomography

All brain tissue included in this study was processed in accordance with previously published methods^2^. At autopsy, fresh samples were collected and dissected into blocks of cortical tissue at dimensions of approximately 1 x 1 x 5 mm containing all six layers of the neocortical mantle. For downstream array tomography, dissected tissue was first immersed in AT fixative (10 mL 16% PFA + 1.0g sucrose + 4mL 10X PBS + 26.0mL dH_2_O) for a period of 2-3 hours and washed in AT wash buffer (5.0mL 10X PBS + 1.25g sucrose + 0.188g glycine + 45.0mL dH_2_O) for five minutes at room temperature. Dehydration was then performed by placing tissue in ascending grades of ethanol (50%, 70%, 95% and twice at 100%) for five minutes per wash. Dehydrated tissue was infiltrated with resin via immersion in 50% LR white in ethanol (Agar Scientific: AGR1281A) before transferal to 100% LR white, both for five minutes each. Tissue was then incubated in a second change of fresh 100% LR white overnight at 4°C. The following day, tissue was placed in gelatin capsules filled with 100% and orientated such that all six cortical layers were approximately parallel to the eventual cutting plane for production of AT ribbons. Resin was then allowed to cure at 56°C for 48-72 hours or until otherwise confirmed to have set fully. Tissue blocks were stored at room temperature until required for experimentation.

### Tissue Sectioning for Array Tomography and Immunogold EM

Tissue was sliced into ultrathin sections for either array tomography or immunogold EM using a Leica UC7 ultramicrotome (Lieca Camera; Wetzlar, Germany. For array tomography, resin blocks from BA20/21 were first faced using a DiATOME Trim 45 blade (TT-45) before ribbons of contiguous 70nm-thick slices were produced using a DiATOME Histo Jumbo diamond blade (80-HISJ). Once sectioned, ribbons were annealed to 22 x 40mm 100pcs 1.5 thickness borosilicate coverslips (VWR: 631-0136) pre-treated with a filtered gel solution (0.1% gelatin from cold water fish skin + 0.01% chromium potassium sulphate in dH2O). Once annealed to the gel-treated coverslips, ribbons were allowed to dry before being stored until required for further experimentation.

For immunogold EM, resin-embedded tissue blocks, processed for AT, were faced as described above before being sliced into 50nm ultrathin sections. Up to 2-3 sections were mounted at a time on to formvar/carbon-coated nickel EM grids (Agar Scientific: S162N7) and allowed to dry for 3-5 minutes. Grids were then stored at room temperature until required.

### Immunostaining of Human Tissue

#### Immunostaining of Array Tomography Ribbons

Tissue ribbons were washed for five cycles with TBS (Fisher Bioreagents: BP2471) via continuous flow. Following initial washing, ribbons were then incubated for 10 minutes in 100µL 50mM glycine solution. Glycine solution was then eluted and ribbons washed for a further five cycles in TBS. Blocking of non-specific antibody binding was performed via incubation of ribbons in 100µL syringe-filtered AT blocking reagent (0.1% fish skin gelatin + 0.05% TWEEN^Ò^20 in TBS) for 30 minutes at room temperature. Ribbons were then washed for a further five cycles in TBS and 100µL primary antibodies (**Supplementary Table S1**) added and incubated overnight at 4°C. The next day, ribbons were washed for five cycles with TBS and incubated with 100µL secondary antibodies (**Supplementary Table S1**) for 30 minutes at room temperature, protected from light. Secondary antibodies were eluted by washing ribbons for a final five cycles in TBS before mounting on to 25 x 75 x 1mm SuperFrost^TM^ Plus Adhesion microscope slides (Epredia: J1800AMNZ) using aqueous hardcuring Immu-Mount^TM^ mounting medium (Fisher Scientific: 10662815). Slides were sealed using clear nail varnish and stored at 4°C, protected from light, until required for imaging but for no longer than 14 days to avoid degradation of fluorescent signal intensity. Ribbons were stained in discrete batches and, to limit potential bias, each batch was balanced to contain a mixture of samples sourced from both Control and AD experimental groups.

### Immunogold Labelling of Ultrathin Sections for Electron Microscopy

Immunogold staining was conducted in accordance with methods detailed previously^2,3^. Briefly, formvar/carbon coated nickel grids (Agar Scientific: S162N7) were incubated first in a saturated solution of sodium metaperiodate in boiled dH_2_O for one minute, followed by five minutes in 1% sodium borohydride. Grids were then incubated in 50mM glycine in 0.1M PB for 10 minutes before blocking for 1 hour at room temperature (0.1% BSA + 0.1% fish skin gelatin + 0.05% TWEEN^Ò^20 in 0.2M PB; pH 7.4). Lecanemab primary antibody was diluted to a working concentration of 1/50 in immunogold EM dilution buffer (0.1% BSA + 150mM NaCl in 0.2M PB) and grids incubated with this primary antibody solution overnight at 4°C. The next day, grids were washed in six changes of 0.2M PB for two minutes per wash. Secondary antibody solution was then formulated by diluting goat anti-human IgG (H+L) 10nm gold stock antibody solution (Abcam: ab39596) to a working concentration of 1/50 in immunogold EM dilution buffer. Grids were incubated in secondary antibodies at room temperature for 90 minutes before washing in six changes of 0.2M PB again for two minutes per wash. Post-staining was performed by incubating grids in 0.1% osmium tetroxide for 20 minutes at room temperature before washing in three cycles of boiled dH_2_O for two minutes, each. Negative staining was then carried out by incubating grids in 3% uranyl acetate in 50% ethanol for 15 minutes before rinsing for three cycles by dipping in dH_2_O. Grids were then stained for 2.5 minutes with lead citrate buffer (4.3% lead nitrate + 5.9% sodium citrate + 2.7% 10M sodium hydroxide in dH_2_O, pH 12.0). A final three washes were then performed by dipping grids in dH_2_O as described previously and grids subsequently stored at room temperature until required for imaging.

### Image Acquisition

#### Imaging of Immunostained Array Tomography Ribbons

Serial images of ribbons were acquired using a Zeiss Axio Imager Z2 epifluorescent microscope, fitted with a CoolSnap digital camera (Zeiss, Baden-Württemburg, Germany). Zen Blue 3.0 software was used to acquire images using a 63X 1.4 NA Plan Apochromat objective. A single tissue landmark (e.g. a plaque, blood vessel or vacuole) was identified visually and used to generate serial images along the length of each ribbon at the same coordinate. Raw image series were obtained in multi-channel 16-bit greyscale (1392 x 1040p) format and saved as .czi files. To avoid bias and ensure comparability, image acquisition settings were calibrated to a single positive control for all stains at the beginning of each imaging session (corresponding to a single immunostaining batch) and identical settings then used to image all other samples within said session.

For each sample, a minimum of three image series were acquired from two resin-embedded tissue blocks; amounting to a total of six image series per sample (or three for samples that had only one resin block available). Where plaques were present, up to three stacks were ideally acquired with a plaque centered in the imaging field, while the remaining three were absent of any plaques. Sampling of imaging fields was conducted randomly across all six cortical layers and prioritized dense core over diffuse plaques. Where no dense core plaques were present, diffuse plaques were instead sampled. To provide unbiased confirmation of plaque morphology, all acquired images of plaques (N = 84) were classified as either “dense core”, “diffuse” or “atypical” by two independent experimenters who were blinded to experimental group. Of the plaques assessed, those classified as “atypical” (N = 3) were excluded from all analysis. Furthermore, a number of samples did not possess the full complement of two resinembedded tissue blocks, or, due to heterogeneity in plaque load, presented with either less than three or no plaques within the excavated volume of tissue. As mentioned, these cases had only three image stacks acquired from a single resin-embedded block and were still included for analysis. The full complement of cases, alongside the number of resin-embedded blocks available and stacks acquired are detailed in **Supplementary Table S2**.

### Image Analysis

#### Stacking, Processing and Analysis of Array Tomography Serial Images

Image series were stacked into 3D volumes using in-house Fiji macros. Proprietary MATLAB scripts were then used to align stacks via rigid and affine registration. Damaged, misaligned or debris-filled sections were removed manually from either end of the stack and images cropped centrally to remove out-of-focus regions at the edges. Cropped stacks were then processed via median filter (pixel radius = 1px) and rolling ball background subtraction. Background subtraction parameters were set based on the size of objects within each channel, with synaptophysin and PSD95 processed with a smaller radius of 10.0px, lecanemab processed with a radius of 100.0px and GFAP, 70.0px. To limit bias, median filter and background subtraction parameters for each channel were kept constant across all images.

Processed stacks were then blinded using an external Fiji plugin (https://github.com/imagejan/blind-experiment) and, using MATLAB scripts, segmented via auto-local thresholding to produce binary images. Only fluorescent objects tracking through at least two consecutive sections of the stack were retained as genuine 3D objects, with any failing to track through automatically filtered out by the segmentation algorithm as non-specific noise. Parameters for segmentation were selected based on their ability to reproduce objects within the original greyscale image and, to limit bias, identical parameters were used to segment all image stacks within a given staining batch.

Due to a rare spectral bleed-through observed from GFAP (AF647) into the synaptophysin/SY38 channel (AF594), segmented images were further processed prior to data extraction to avoid confounds arising from this technical artefact. For all image stacks, synaptophysin and GFAP channels were first overlaid in Fiji and the ‘Image Calculator’ utility used to subtract any overlapping GFAP signal from the former. By performing this additional processing step, potential effects of GFAP signal bleed through into the presynaptic channel were effectively masked out.

Data were extracted from segmented images as either object density or colocalisation measures. For density, a custom MATLAB script was used to generate a 3D neuropil mask using synaptophysin as the reference channel to exclude regions of negative synaptic staining, such as blood vessels. The number of objects within the volume of each neuropil mask was then automatically calculated for each image stack and scaled mathematically to be expressed in terms of density of objects per mm^3^ of tissue. For colocalisation, an in-house Python script, distributed via Docker container (client version 24.0.7) was used for data extraction. Pre- and postsynaptic objects were registered as a “synaptic pair” by the script only when corresponding object centroids resided within a 0.5µm distance. For colocalisation between paired synapses and lecanemab, at least 50% of the synaptic object volume was required to overlap with a corresponding lecanemab-positive 3D object. Colocalisation of synapses with lecanemab was expressed quantitatively as the relative percentage (%) of total paired pre- or postsynaptic objects found to successfully overlap with lecanemab.

For quantification of synapse density and colocalisation with lecanemab as a function of plaque distance, only dense core plaques from the AD group with an agreed classification between both blinded experts were included for analysis. Image stacks passing initial inclusion criteria that further showed clear evidence of a second plaque within the imaging field (N = 4) were excluded, amounting to the final numbers reported for analysis here (N = 40 plaques across N = 18 AD cases). Greyscale image stacks corresponding to each plaque were converted to maximum intensity projections, blinded via Fiji plugin (https://github.com/imagejan/blindexperiment) and ROIs then drawn around central cores of the blinded plaques using the “polygonal selection” tool in Fiji. After unblinding, ROIs were then applied to the corresponding segmented versions of the same image stacks and the “clear outside” function applied to delineate the segmented core from surrounding signal. Using an in-house MATLAB script, segmented stacks containing total paired synapses and paired pre/post synapses with lecanemab were binned into concentric 10µm radii from the isolated plaque core boundary, extending to a maximum distance of 50µm. Within each individual bin, the number of paired synaptic objects (total or colocalised with lecanemab) were then quantified. Within each bin, the density or percentage (%) of paired pre/postsynapses with lecanemab were then calculated.

### Statistical Analysis

All statistical analysis was carried out using *R* (Version 4.2.1, June 2022). Data in raw format were imported into R as .csv files and encoded as either, integers, floating point numbers (decimals) or categorical factors. Once imported, data were first assessed for normality through visual inspection of histograms in conjunction with formal Shapiro-Wilk testing. A ShapiroWilk *p < 0.05* was taken to indicate non-normally distributed data. Based on the outcomes of normality assessment, data were then collapsed into either single mean datapoints per sample if normally distributed, or median datapoints in the case of a skewed distribution. Averaged datapoints are used only for illustrative purposes in graphs (excepting **Figure 3B-D**, described further below), while the full (un-averaged) distribution of data was used for downstream linear mixed effects analysis.

Analysis of data for **Figures 1**, **2** and **Supplementary Figures S1-S4/S6-S7** utilized linear mixed effects models (LMMs) adjusting for sex, APOE4 status and PMI as fixed effect covariates. Tissue block nested within case ID was fitted as the random effect term to account for correlated within-case and within-block variance; modelling the full distributional properties of the data without pseudoreplication and thus negating the need to collapse said data down into single averages. To further exclude the possibility of overfitting, LMM fit was assessed via comparison of Akaike Information Criterion (AIC) and Bayesian Information Criterion (BIC) values between increasingly complex model structures, with lower AIC/BIC values taken to indicate more optimal fit.

For both standard linear models and LMMs, assumptions were tested after establishing final model structures. First, absence of multicollinearity was established by calculating variance inflation factors (VIFs) for each variable. A VIF < 5.0 was taken to indicate low/absent correlation between model covariates. Second, models were tested for normality of residuals through visual inspection of residual and QQ plots, combined with formal Shapiro-Wilk normality testing. Although Shapiro-Wilk *p < 0.05* was taken to indicate a lack of normally distributed residuals, visual inspection was the prime method for determining whether model residuals approximated toward a normal distribution. Finally, homoscedasticity was tested via Levene’s test, with *p < 0.05* taken to indicate unequal variance. For models found to meet assumptions, the significance of main effects were assessed by extracting p-values for each variable via Type III ANOVA with Satterthwaite approximation for degrees of freedom. Models failing to meet assumptions were re-constructed following transformation of the response variable via Tukey Ladders of Power prior to derivation of p-values using the same method described above to ascertain significance of main effects. In both cases, *p < 0.05* for any variable was taken to indicate a statistically significant main effect. For analysis of variables in the form of factors with >2 levels, post-hoc pairwise comparisons were performed using the *emmeans()* function with Holm’s correction for multiple testing; *p < 0.05* being interpreted as a significant pairwise difference between any two groups.

For analysis regarding the assessment of paired synapse density and colocalisation of pre/postsynapses with lecanemab as a function of plaque distance (**Figure 3B-D**), linear mixed effects models could not be employed for analysis. This was due to the fact that, after filtering data to include only dense core plaques that had reached an agreed classification by both blinded experts, three cases had only one plaque remaining that had passed inclusion criteria and hence, within-subject variance could not be estimated for this data. With this in mind, analysis of normally distributed paired synapse density as a function of plaque distance was performed on averaged (mean) datapoints for each sample via simple linear models adjusting for sex, APOE4 status and PMI. Linear models for each bin were constructed to compare the paired synaptic density against the baseline of the control group—defined as the mean of image stacks from said control group with no plaque present. On the other hand, data relating to the % of surviving paired pre/postsynapses within each bin found to colocalise with lecanemab, which was not normally distributed, was instead performed through Wilcoxon Rank Sum test, comparing each bin against the median of image stacks from the control groups with no plaque present. In both cases, *p-values* for each bin were manually corrected for five comparisons (for each bin extending from 10-50µm) and these adjusted p-values were used to interpret significant difference from the control baseline, as illustrated in the main figure.

## Supplementary Figures and Tables

### Supplementary Figures

**Supplementary Figure S1:**
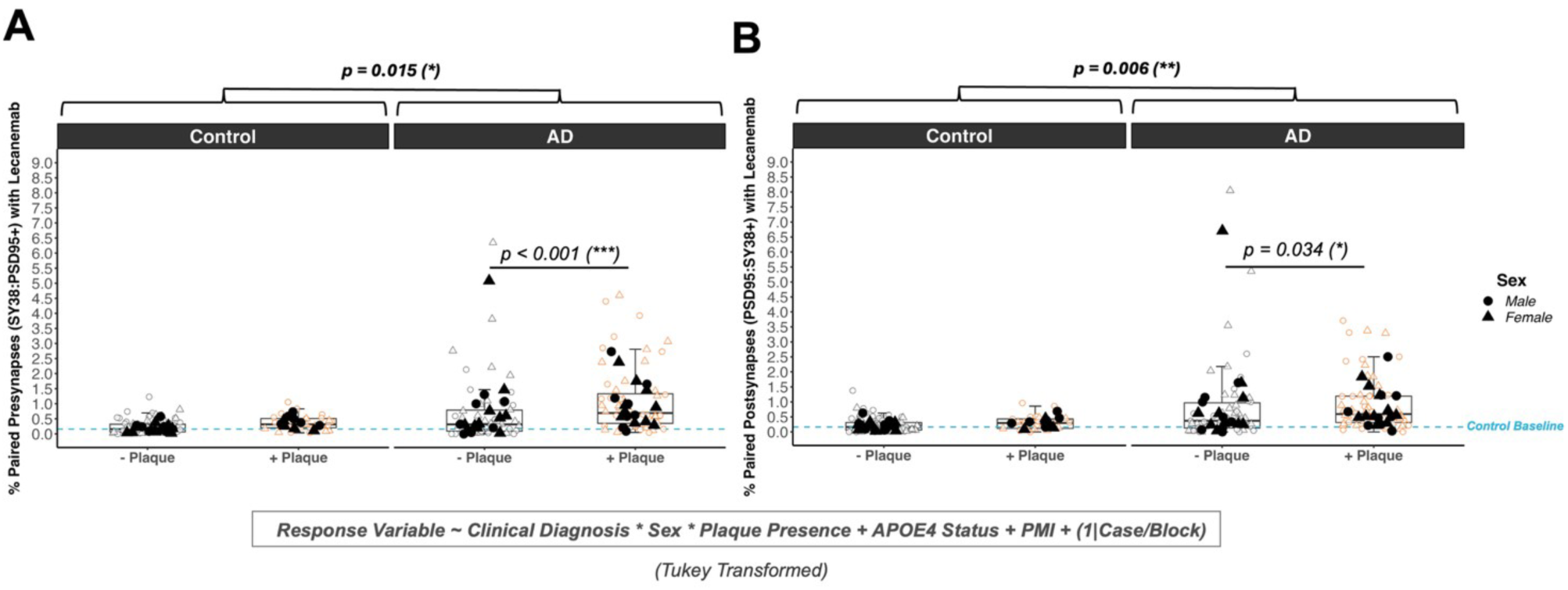
Pre- and Postsynapses Exhibit Greater Association with Lecanemab in the AD Temporal Cortex when Residing Close to Plaques. (**A**) Quantification of the percentage (%) of total paired presynaptic objects colocalised with lecanemab after further stratifying data by plaque presence. After adjusting for plaque presence, AD Cases (*N* = 20) were still observed to exhibit globally elevated colocalisation over controls (*N* = 19). In AD, the presence of a plaque was however further associated with a significantly greater likelihood of observing lecanemab colocalised with presynaptic puncta when compared against plaque distal imaging fields. (**B**) Quantification of paired postsynaptic objects colocalised with lecanemab exhibited a similar pattern of data. Filled datapoints indicate average (median) paired pre- or postsynaptic objects colocalised with lecanemab per human case, while clear datapoints indicate underlying technical replicates in the form of individual image stacks (*N* = 3-6 stacks per case). **Abbreviations**: AD = Alzheimer’s Disease.

**Supplementary Figure S2:**
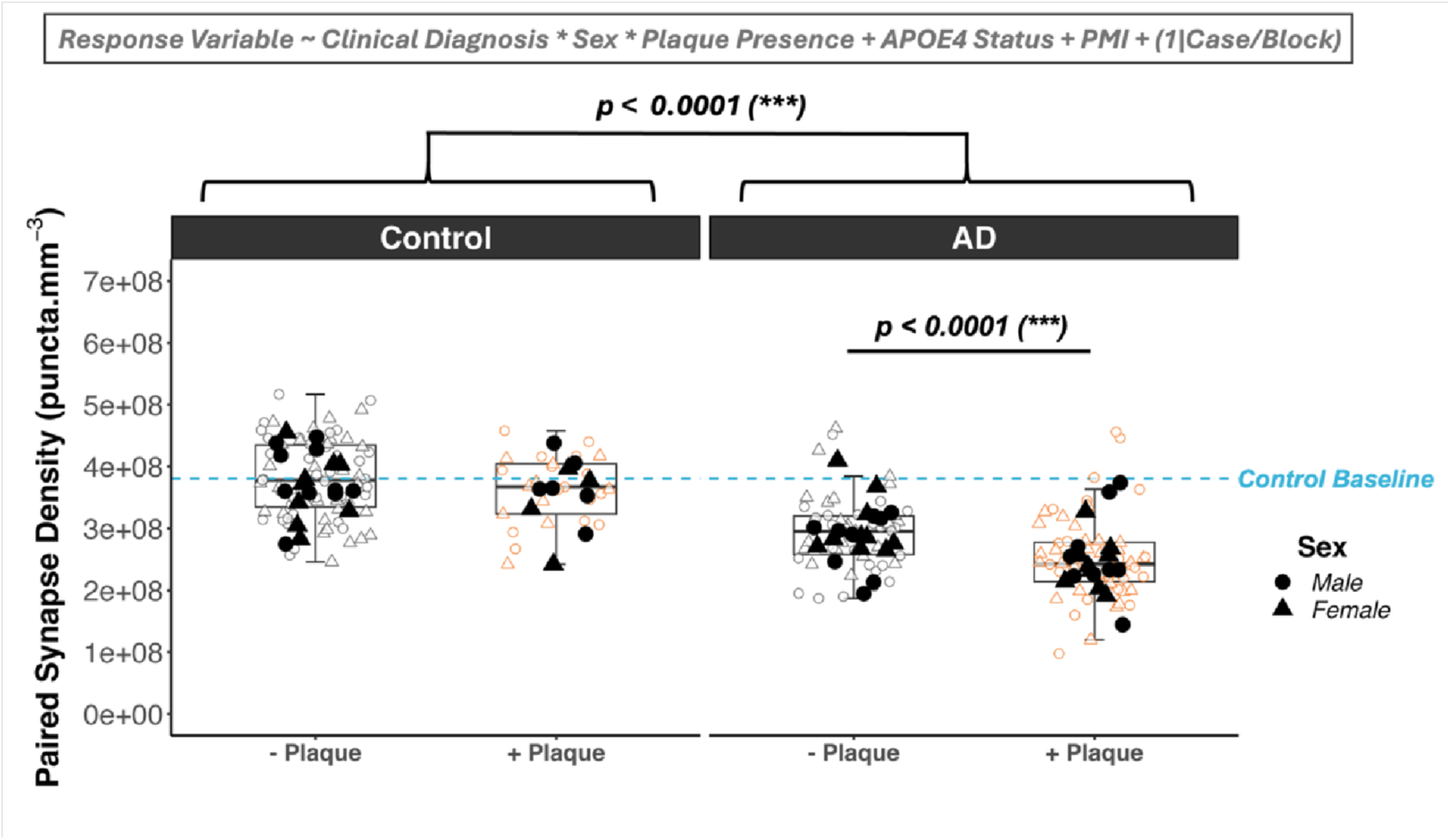
Paired Synapse Density is Further Reduced in Association with Dense Core Plaques in the AD Temporal Cortex. Quantification of paired synapse density, stratified by plaque presence (both dense core and diffuse) revealed a significant overall loss of synapses in AD cases below the control baseline, represented by the mean of control image stacks with no plaque present (blue dashed line). This synapse loss was observed even in plaque distal regions but was further exacerbated by the presence of plaques. Filled datapoints indicate average (mean) paired synapse density per human case, while clear datapoints indicate underlying technical replicates in the form of individual image stacks (N = 3-6 per case). **Abbreviations:** AD = Alzheimer’s Disease.

### Supplementary Tables

**Supplementary Table S1:** (**A**) Primary antibodies used for either AT or iEM. (**B**) Corresponding Secondary Antibodies used for AT or iEM. **Abbreviations**: AT = Array Tomography, iEM = Immunogold Electron Microscopy.

| A — PRIMARY ANTIBODIES |  |  |  |  |  |  |  |  |
| --- | --- | --- | --- | --- | --- | --- | --- | --- |
| Name | Used in: | Host Species | Dilution | Working Conc. (ug/mL) | Supplier | Cat. ID | RRID | Lot No(s). |
| Anti-Synaptophysin (SY38) | AT | Mouse | 1/50 | 1.0 | Abcam | ab8049 | AB_2198854 | 1096179-1 |
| Anti-PSD95 (D27E11) | AT | Rabbit | 1/50 | 0.38 | Cell Signaling Technology | 3450 | AB_2292883 | Lot: 5 |
| Lecanemab | AT, iEM | Human | 1/400 (AT),<br>1/50 (iEM) | 1.0 (AT),<br>14.0 (iEM) | Provided Courtesy of Prof. Bart De Strooper | — | — | — |
| B — SECONDARY ANTIBODIES |  |  |  |  |  |  |  |  |
| Name | Used in: | Host Species | Dilution | Working Conc. (ug/mL) | Supplier | Cat. ID | RRID | Lot No. |
| *Donkey Anti-Mouse IgG (H+L) Alexa Fluor 594* | AT | Donkey | 1/50 | 40.0 | Invitrogen | A21203 | AB_2535789 | 2474956 |
| *Donkey Anti-Human IgG (H+L) DyLight 405* | AT | Donkey | 1/50 | 30.0 | Jackson ImmunoResearch | 709-475149 | AB_2340553 | 171546 |
| *Donkey Anti-Rabbit IgG (H+L) Alexa Fluor 488* | AT | Donkey | 1/50 | 40.0 | Invitrogen | A21206 | AB_2535792 | 2541645 |
| *Goat Anti-Human IgG (H+L) 10nm Gold* | iEM | Goat | 1/25 | — | Abcam | ab39596 | AB_954439 | 1080942-1 |

**Supplementary Table S2:** Limited availability of resin blocks for a small number of cases meant that only 3 image stacks could be acquired from a single block. Furthermore, heterogeneity in plaque load across experimental groups meant that, while a maximum of 3 plaques could be sampled from the majority of AD cases, a number of cases from the control group yielded < 3 (N = 13) after sectioning of tissue at the scale required for AT. **Abbreviations:** AT = Array Tomography.

| Case BBN No. | Clinical Diagnosis | Resin Blocks Available | Total No. Image Stacks | No. Plaques Sampled |
| --- | --- | --- | --- | --- |
| BBN001.28406 | Control | 2 | 6 | 2 |
| BBN001.32577 | Control | 2 | 6 | 0 |
| BBN001.35181 | Control | 2 | 6 | 3 |
| BBN001.35215 | Control | 2 | 6 | 0 |
| BBN001.36135 | Control | 1 | 3 | 1 |
| BBN001.36435 | Control | 2 | 6 | 2 |
| BBN001.37786 | Control | 2 | 6 | 3 |
| BBN001.28793 | Control | 2 | 6 | 0 |
| BBN001.28794 | Control | 1 | 3 | 0 |
| BBN001.36809 | Control | 2 | 6 | 0 |
| BBN001.26495 | Control | 2 | 6 | 1 |
| BBN001.28797 | Control | 2 | 6 | 3 |
| BBN001.34131 | Control | 2 | 6 | 3 |
| BBN001.29082 | Control | 2 | 6 | 3 |
| BBN001.29086 | Control | 1 | 3 | 0 |
| BBN001.37555 | Control | 2 | 6 | 0 |
| BBN001.35866 | Control | 2 | 6 | 3 |
| BBN001.28402 | Control | 2 | 6 | 2 |
| BBN_19686 | Control | 2 | 6 | 0 |
| BBN001.29695 | AD | 2 | 6 | 3 |
| BBN001.35182 | AD | 2 | 6 | 3 |
| BBN001.37711 | AD | 2 | 6 | 3 |
| BBN001.28771 | AD | 2 | 6 | 3 |
| BBN001.32929 | AD | 1 | 3 | 3 |
| BBN_25739 | AD | 2 | 6 | 3 |
| BBN001.38151 | AD | 2 | 6 | 2 |
| BBN001.30142 | AD | 2 | 6 | 3 |
| BBN001.36297 | AD | 2 | 6 | 3 |
| BBN001.37400 | AD | 2 | 6 | 3 |
| BBN001.36346 | AD | 2 | 6 | 3 |
| BBN001.35535 | AD | 2 | 6 | 3 |
| BBN001.29081 | AD | 2 | 6 | 3 |
| BBN001.30853 | AD | 2 | 6 | 3 |
| BBN001.35564 | AD | 2 | 6 | 3 |
| BBN001.30883 | AD | 2 | 6 | 3 |
| BBN001.30973 | AD | 2 | 6 | 3 |
| BBN_24322 | AD | 2 | 6 | 3 |
| BBN_24526 | AD | 2 | 6 | 3 |
| BBN_24527 | AD | 2 | 6 | 2 |

